# Knowledge, attitude, and practice regarding type 2 diabetes and associated factors among rural adolescents in Indonesia: A cross-sectional study

**DOI:** 10.1101/2025.09.25.25336672

**Authors:** Nila Kusumawati, Hanno Pijl, Abdul Hamid, Emul Yani, Desti Puswati, Halimatou Alaofè

**Author notes:** Corresponding Author: (NK).

## Abstract

**Background:** Type 2 diabetes (T2D) is increasingly affecting adolescents globally, yet there is limited evidence on their knowledge, attitudes, and practices (KAP) regarding the disease, which is essential for promoting preventive strategies. This study aims to evaluate KAP levels among adolescents, particularly in rural Indonesia, and identify factors associated with these outcomes.

**Methods:** A cross-sectional study was conducted in Kampar Regency, Riau Province, Indonesia, from September 2024 to February 2025. The study involved 1,546 senior high school students, who were selected through multistage cluster sampling. Data were collected using a validated KAP questionnaire. Descriptive statistics were used to summarize the levels of KAP, while multivariable binary logistic regression identified associated factors.

**Results:** About 70.6%, 70.3%, and 74.6% of participants showed good knowledge, positive attitudes, and good practices related to T2D. Higher likelihoods of good knowledge were linked to being female (AOR = 1.44; 95% CI: 1.13–1.83), non-Indigenous (AOR = 1.78; 95% CI: 1.40–2.28), studying in peri-urban areas (AOR = 1.43; 95% CI: 1.11–1.84), having longer school hours (AOR = 1.68; 95% CI: 1.07–2.63), being in higher grades (AOR = 1.51; 95% CI: 1.20–1.90), ranking in the top ten (AOR = 1.60; 95% CI: 1.24–2.05), participating in extracurriculars (AOR = 1.49; 95% CI: 1.16–1.92), and having a father with higher education (AOR = 1.32; 95% CI: 1.01–1.72). Positive attitudes and good practices were more common among students from periurban areas (AOR = 1.55; 95% CI: 1.20–2.00), those with longer school hours (AOR = 1.92; 95% CI: 1.22–3.02), higher grades (AOR = 1.33; 95% CI: 1.06–1.68), and top ten rankings (AOR = 1.63; 95% CI: 1.27–2.10). Additionally, adolescents aware of first-degree relatives’ T2D status were more likely to show positive attitudes (AOR = 0.70; 95% CI: 0.53–0.92) and good practices (AOR = 0.72; 95% CI: 0.54–0.96).

**Conclusion:** Adolescents in rural Indonesia demonstrated good knowledge, attitude, and practices related to T2D, but notable gaps remain across all domains. Targeted interventions in schools that consider significantly associated factors, along with active parental involvement in promoting healthy habits at home, are crucial for addressing these gaps and preventing future T2M burden in the country.

## Introduction

Type 2 diabetes (T2D) is a significant non-communicable disease in Indonesia, with increasing rates of morbidity and mortality [1,2]. According to the latest *Diabetes Atlas* by the International Diabetes Federation (2025), Indonesia ranks fifth globally in diabetes prevalence among individuals aged 20 to 79, and it is third in terms of undiagnosed cases. Projections indicate that the number of people affected by diabetes and its complications will continue to rise [1].

Moreover, diabetes has been identified as the fifth leading cause of death among non-communicable diseases in the country [3]. A 2016 study revealed that 4% of the 18.9 million enrollees in Indonesia’s National Health Insurance were patients with T2D, with 57% of these individuals experiencing complications. The direct medical costs associated with diabetes amounted to USD 576 million, with 74% of this budget dedicated to managing complications [4].

Type 2 diabetes is increasingly being diagnosed in adolescents, despite its historical perception as an adult-onset disease [5, 6]. Risk factors such as obesity, physical inactivity, and smoking are significant contributors to this trend, especially in Indonesia, where a low diagnosed prevalence (less than 0.05%) may not reflect the true burden due to many undiagnosed cases [7, 8]. The COVID-19 pandemic has led to a surge in gadget use among adolescents, resulting in excessive screen time that correlates with decreased physical activity and increased body mass index (BMI) [9–11]. This growing reliance on digital devices has adversely affected healthy lifestyle habits, impacting adolescents in both urban and rural areas of Indonesia, where internet access has significantly expanded [12–14].

Therefore, reducing adolescents’ exposure to risk factors for T2D is crucial for diminishing its future prevalence in Indonesia. A study suggests that with effective prevention programs, diabetes rates could drop to 9.22% [15]. A key first step is assessing adolescents’ knowledge, attitude, and practice (KAP) regarding T2DM. However, research indicates significant disparities in KAP levels in high-burden countries across different demographics; for example, students in peri-urban Bangladesh showed more knowledge than those in rural areas [16], and Nepalese metropolitan students had higher knowledge than their rural peers, despite similar attitudes [17]. In Jordan, urban adolescents had moderate to high knowledge, but their attitudes and practices were lacking [18]. A study in India found that rural high school students had a fair understanding of diabetes, but their attitudes and self-care practices were not assessed [19].

Additionally, research on KAP concerning T2D in rural settings, particularly in light of lifestyle changes after COVID-19, is limited. This study aims to assess KAP levels among rural Indonesian students and identify associated sociodemographic factors. Understanding these dynamics is vital for guiding health policymakers in Indonesia to develop targeted and culturally relevant interventions. Raising awareness about diabetes at a young age can facilitate early identification and reduce the disease’s severe consequences. Furthermore, these findings could be beneficial for other low- and middle-income countries, which are projected to see a 95% rise in diabetes prevalence by 2050 [1]. Generating context-specific evidence is essential for shaping effective local and global strategies to address the growing burden of T2D.

## Materials and Methods

### Study design, setting, and period

This study was conducted from September 18, 2024 to February 7, 2025 in Kampar Regency, Riau Province, Indonesia. It used a cross-sectional descriptive design to assess KAP levels at a specific point in time [20]. According to the latest data from the Indonesian Health Survey, Riau Province ranks 15th out of 38 provinces in terms of diabetes prevalence, with a rate of 1.5%, which is slightly lower than the national average. Unhealthy lifestyle habits, such as the consumption of sugary drinks and low levels of physical activity, have been observed in children as young as three years old [7–8, 12, 21]. Kampar Regency was selected for this study due to its high diabetes prevalence and the significant number of adolescents in senior high schools [21].

### Study population, sample size, and sampling

The study targeted 21,872 senior high school students in Kampar Regency. Using OpenEpi (version 3) for a cross-sectional design, we calculated a sample size of 1,288 students at a 99% confidence level, with an anticipated percent frequency of 50%, 5% confidence limits, and a design effect of 2 to account for cluster sampling. However, after adjusting for a 20% non-response rate [22], we recruited 1,546 students for better statistical power and data reliability. We employed a multistage cluster sampling technique to facilitate efficient participant selection [23]. First, we randomly selected 11 out of 21 districts. Next, one senior high school was chosen from each selected district, and finally, students were recruited from these schools using a proportional allocation method. Inclusion criteria required voluntary participation and attendance during the study period.

### Data collection instrument and procedure

Data collection was conducted using a pretested questionnaire adapted from relevant diabetes literature [16]. This questionnaire was refined with input from experts in various health fields, including Internal Medicine, Endocrinology, Diabetology, Nursing, and Public Health. Two researchers evaluated the items for face validity, followed by an assessment of content validity by six experts. This process resulted in an Item-Level Content Validity Index (I-CVI) of 0.80 and a Scale-Level Content Validity Index (S-CVI) of 0.90. The questionnaire was then pilot-tested on 420 participants from three public senior high schools in Kampar Regency, ensuring that these participants were not included in the main study. Validity was confirmed through Pearson correlation analysis, with correlation coefficients ranging from 0.273 to 0.585, exceeding the threshold necessary to confirm their validity in measuring the intended constructs. Construct validity was assessed using the Kaiser-Meyer-Olkin (KMO) measure, which yielded a KMO value of 0.779, and Bartlett’s Test, which showed a significance level of p < 0.001. These results indicate that the data are suitable for exploratory factor analysis. Lastly, internal consistency was confirmed with a Cronbach’s Alpha of 0.725, suggesting acceptable reliability.

The questionnaire, conducted in Indonesian, comprised four sections. The first section collected socio-demographic information, including age, gender, ethnicity, grade level, class rank, school attendance, participation in extracurricular activities, parents’ education and employment status, family history of diabetes, and height and weight. To measure height, the participant stands barefoot against a wall or stadiometer, lowering the headpiece to the crown of their head. For weight, the child stands on a digital scale in the center, removing shoes and heavy clothing.

Record height to 0.1 cm and weight to the nearest decimal. Height and weight were classified into obese and non-obese categories based on the Indonesian BMI classification using WHO cutoffs [24]. The second section assessed participants’ knowledge about diabetes with five questions. Three allowed for multiple correct answers, while two used ‘yes,’ ‘no,’ and ‘do not know’ options. Participants earned 1 point for each correct answer. The third section evaluated attitudes with six questions on diabetes prevention and risk factors. Positive responses received 1 point, while negative and ‘do not know’ responses scored 0. The final section assessed practices related to physical activity, smoking, and weight monitoring, with scores ranging from 0 (least healthy) to a maximum indicating the healthiest behavior. Overall, the total KAP score for diabetes ranges from 0 to 32, with knowledge scores from 0 to 18, attitude scores from 0 to 6, and practice scores from 0 to 8. Data were collected in school classrooms using a self-administered questionnaire, with assistance from trained students in Public Health, Nursing, Nutrition and Midwifery, under the supervision of the principal investigator and co-authors at each school.

### Ethics approval and consent to participate

Ethical approval for the study was obtained from the Institutional Review Board of Institut Kesehatan Payung Negeri Pekanbaru in Pekanbaru, Riau Province, Indonesia (No. 277/IKES PN/KEPK/IX/2024) on September 3, 2024. Permission to collect data in the participating schools was granted by the Provincial Health Department (No. 421/cabdisdik/6.2/2023/083). Schools and participants were informed about the purpose and procedures of the study, and participants were assured of their voluntary participation and their right to withdraw at any time without penalty.

Prior to data collection, participants aged 17 years and older provided written informed consent, which was witnessed by their homeroom teachers. For participants under 17 years of age, written informed consent was obtained from their parents or legal guardian, and the participants themselves provided assent after the study procedures were explained to them. During data collection, the questionnaire was anonymous, and neither the researchers nor the student assistants present had access to any information that could identify individual participants. After data collection, each completed questionnaire was assigned a unique code to facilitate data entry. The codes did not contain any information that could identify individual participants, ensuring confidentially was maintained. The questionnaires were kept in a locked cabinet in a secure room, which could be only accessed by the researchers.

### Statistical analysis

Descriptive statistics were used to summarize participants’ sociodemographic characteristics and their KAP levels regarding T2D. As each KAP score was non-normally distributed, we categorized them using a median split to simplify interpretation: scores equal to or above the median indicated good knowledge, a positive attitude, or good practice, while those below the median were classified as poor or negative. Then, bivariate analyses examined the relationships between KAP levels and sociodemographic variables, including gender, ethnicity, BMI, grade level, class rank, extracurricular participation, parental education and employment, family diabetes history, and information-seeking behavior, using the Chi-square test. Age was analyzed with the Mann-Whitney U test due to its continuous and non-normally distributed nature.

Variables with p ≤ 0.20 were included in the multivariable model. Finally, to identify sociodemographic factors independently associated with participants’ diabetes-related KAP levels, a multivariable binary logistic regression was conducted. All relevant assumptions were assessed and met. Specifically, Multicollinearity was evaluated using the Variance Inflation Factor, with values ranging from 1.0 to 2.7, indicating no issues. The model fit was confirmed with the Hosmer-Lemeshow goodness-of-fit test (χ²(8) = 10.23, p = 0.249). Statistical significance was set at p < 0.05, with results presented as adjusted odds ratios (AOR) and 95% confidence intervals (CI).

## Results

### Study participants’ sociodemographic characteristics

A total of 1,546 adolescent students with a mean age of 16 years participated in the study. Of these, 64% were female, and 91.8% were non-obese, as shown in Table 1. Most attended periurban schools (63.8%), spent over 8 hours at school daily (93.3%), and 65.8% did not engage in extracurricular activities. Many had parents with higher education levels, with 62.3% of fathers and 61.4% of mothers educated beyond high school. About 19.9% were unsure about diabetes in first-degree relatives, while 9.1% confirmed at least one relative had the disease. Furthermore, 23.5% did not know about T2D, and among those who did, 48.8% cited social media as their main information source.

**Table 1.** Sociodemographic characteristics of the study participants.

| Characteristics | Number | Percentage |
| --- | --- | --- |
| **Age (mean ± SD)** | 16.27 ± 0.783 | |
| 15-19 years | 1,546 | 100 |
| **Gender** |  |  |
| Male | 557 | 36 |
| Female | 989 | 64 |
| **Ethnicity** |  |  |
| Indigenous | 834 | 53.9 |
| Non-indigenous | 712 | 46.1 |
| **Body Mass Index** |  |  |
| Obesity | 126 | 8.2 |
| Non-obesity | 1,420 | 91.8 |
| **School Location** |  |  |
| Peri-urban Area | 987 | 63.8 |
| Remote rural Area | 559 | 36.2 |
| **Daily School Hours** |  |  |
| 8 hours | 103 | 6.7 |
| >8 hours | 1,443 | 93.3 |
| **Class** |  |  |
| Grade X | 716 | 46.3 |
| Grade XI | 830 | 53.7 |
| **Class Rank** |  |  |
| Under-top 10 | 942 | 60.9 |
| Top 10 | 604 | 39.1 |
| **Extracurricular** |  |  |
| Not participated | 1,017 | 65.8 |
| Participated 1 day or more a week | 529 | 34.2 |
| **Father's Employment** |  |  |
| Unemployed | 114 | 7.4 |
| Employed | 1,432 | 92.6 |
| **Mother's Employment** |  |  |
| Unemployed | 1,258 | 81.4 |
| Employed | 288 | 18.6 |
| **Father's Education** |  |  |
| Non-attended or lower levels | 583 | 37.7 |
| Higher educational levels | 963 | 62.3 |
| **Mother's Education** |  |  |
| Non-attended or lower levels | 596 | 38.6 |
| Higher educational levels | 950 | 61.4 |
| **First-degree Relatives' T2D History** |  |  |
| None | 1,098 | 71.0 |
| Yes | 141 | 9.1 |
| No Idea | 307 | 19.9 |
| **Number of First-Degree Relatives**<br>**Diagnosed with T2D** |  |  |
| None | 1,045 | 90.9 |
| One or more | 141 | 9.1 |
| **Having Information about T2D** |  |  |
| Did not receive | 364 | 23.5 |
| Received | 1,182 | 76.5 |
| **Sources of Information about T2D** |  |  |
| School | 403 | 26.1 |
| Television | 366 | 23.7 |
| Website | 656 | 42.3 |
| Social media | 754 | 48.8 |
| Radio | 36 | 2.3 |
| Newspaper | 27 | 1.7 |
| Others (parents, families, neighbours, communities) | 19 | 1.2 |
| **Number of Sources Accessed for Diabetes Information** |  |  |
| 0 source | 364 | 23.5 |
| 1 or more sources | 1,182 | 76.5 |

### T2D-related knowledge

Table 2 shows participants’ responses to knowledge-based questions about T2D. Around 16.6% of participants identified T2D as an infectious disease, and 22.4% were uncertain. Regarding risk factors, 50.8% recognized physical inactivity, while just 8.9% knew smoking increased risk. For symptoms, 53.0% recognized frequent urination, but only 16.9% noted increased thirst and 16.2% excessive hunger. Most (61.5%) knew about foot problems as a complication, but only 13.0% were aware of eye damage risks. Additionally, 15.7% were unsure about preventability, and 2.7% believed T2D was not preventable. The median knowledge score was 5 (IQR 3; range 3-18). Using this median as a cutoff, 1,091 of 1,546 participants (70.6%) were classified as having good knowledge of T2D.

**Table 2.** Participants’ diabetes knowledge (n=1,546).

| Questions and Selected Responses | Frequency | Percentage |
| --- | --- | --- |
| **T2D is an Infectious Disease** |  |  |
| Yes | 256 | 16.6 |
| No | 943 | 61.0 |
| No Idea | 347 | 22.4 |
| **T2D Risk Factors** |  |  |
| Obesity | 603 | 39 |
| Physical inactivity | 785 | 50.8 |
| Family history | 814 | 52.7 |
| Stress | 140 | 9.1 |
| Smoking | 137 | 8.9 |
| **T2D Signs and Symptoms** |  |  |
| Frequent urination | 819 | 53.0 |
| Tiredness | 570 | 36.9 |
| Increased thirst | 262 | 16.9 |
| Increased hunger | 250 | 16.2 |
| Unintended weight loss | 543 | 35.1 |
| **T2D Complications** |  |  |
| Eye blood vessel damage | 201 | 13.0 |
| Kidney damage | 434 | 28.1 |
| Foot problems | 951 | 61.5 |
| Nerve damage | 323 | 20.9 |
| Heart disease | 229 | 14.8 |
| Stroke | 207 | 13.4 |
| **T2D is Preventable** |  |  |
| Yes | 1,262 | 81.6 |
| No | 42 | 2.7 |
| No Idea | 242 | 15.7 |

### T2D-related attitude

As shown in Table 3, 27.7% disagreed that family history increases T2D risk, while 28.5% were unsure. Additionally, 24.4% disagreed about smoking raising T2D risk, and 43% were uncertain. About 24.8% were unsure of obesity’s role in T2D risk. The median attitude score was 4 (IQR 2; range 0-6). Overall, 70.3% of the 1,546 participants had a positive attitude toward T2D.

**Table 3.** Participants’ diabetes attitude (n=1,546).

| Questions and Selected Responses | Frequency | Percentage |
| --- | --- | --- |
| **Healthy Eating is Important to Prevent T2D** |  |  |
| Agree | 1408 | 91.1 |
| Disagree | 19 | 1.2 |
| Not sure | 119 | 7.7 |
| **Exercise Helps Prevent T2D** |  |  |
| Agree | 1378 | 89.1 |
| Disagree | 30 | 1.9 |
| Not sure | 138 | 8.9 |
| **If You Have a Family History of T2D, You Are at Higher Risk of Getting It** |  |  |
| Agree | 677 | 43.8 |
| Disagree | 428 | 27.7 |
| Not sure | 441 | 28.5 |
| **People Who Smoke are More Likely to Get T2D** |  |  |
| Agree | 503 | 32.6 |
| Disagree | 377 | 24.4 |
| Not sure | 655 | 43 |
| **Regular Screening Helps in the Early Detection and Prevention of T2D** |  |  |
| Agree | 1,273 | 82.3 |
| Disagree | 43 | 2.8 |
| Not sure | 230 | 14.9 |
| **Obesity Increases the Risk of Developing T2D** |  |  |
| Agree | 1,034 | 66.9 |
| Disagree | 129 | 8.3 |
| Not sure | 383 | 24.8 |

### T2D-related practice

Table 4 presents the health behaviours of participants and indicates that 16.5% of individuals do not engage in daily physical activity, while 6.4% smoke fewer than 25 cigarettes a day and 2.1% smoke 25 or more cigarettes daily. In terms of weight monitoring, 3.8% of participants never check their weight, 16.0% check their weight annually, and 5.0% monitor it daily. The median practice score was 4 (IQR 2; range 0-8). Out of 1,546 participants, 1,154 (74.6%) demonstrated good practice related to T2D.

**Table 4.** Participants’ diabetes practice (n=1,546).

| Questions and Selected Responses | Frequency | Percentage |
| --- | --- | --- |
| **Daily Physical Activity** |  |  |
| Never | 255 | 16.5 |
| 30 minutes or less | 1021 | 66.0 |
| Less than 60 minutes | 270 | 17.5 |
| **Cigarette Consumption per Day** |  |  |
| Never | 1,414 | 91.5 |
| Less than 25 | 99 | 6.4 |
| 25 or more | 33 | 2.1 |
| **Frequency of Body Weight Monitoring** |  |  |
| Never | 58 | 3.8 |
| Once a year | 248 | 16.0 |
| Once every few months | 540 | 34.9 |
| Once a month | 404 | 26.1 |
| Once a week | 219 | 14.2 |
| Every day | 77 | 5.0 |

### T2D-associated factors

Table 5 highlights factors associated with KAP regarding T2D. Participants showed better knowledge if they were female (AOR = 1.441; 95% CI: 1.133–1.834), non-indigenous (AOR = 1.784; 95% CI: 1.399–2.275), or studied in peri-urban areas (AOR = 1.426; 95% CI: 1.106– 1.838). Increased knowledge was also linked to longer school hours (AOR = 1.676; 95% CI: 1.068–2.630), higher grades (AOR = 1.509; 95% CI: 1.199–1.899), ranking in the top ten (AOR = 1.596; 95% CI: 1.240–2.054), participating in extracurricular activities (AOR = 1.488; 95% CI: 1.155–1.917), and having a father with a higher education (AOR = 1.316; 95% CI: 1.008– 1.719).

**Table 5.**
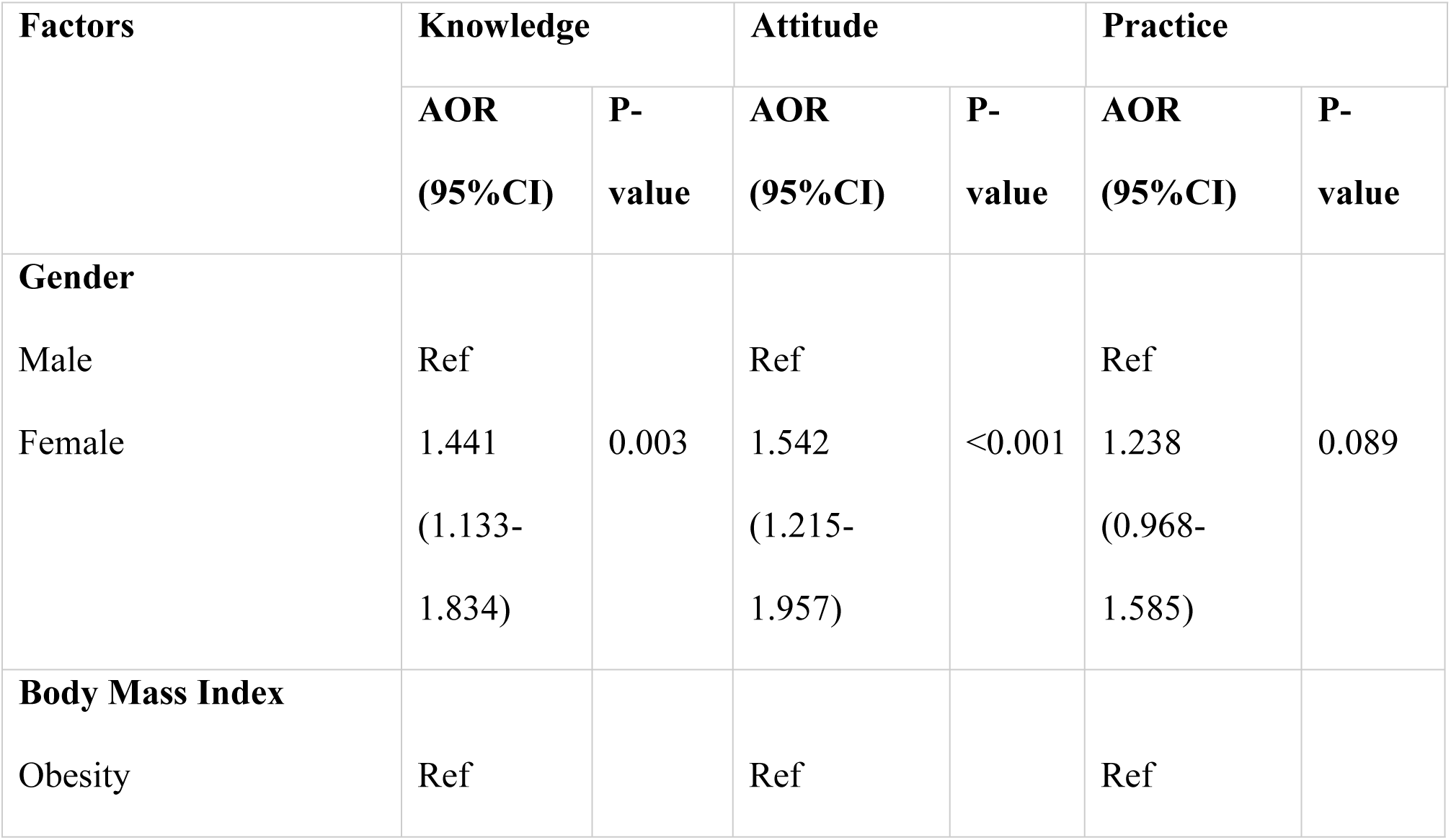
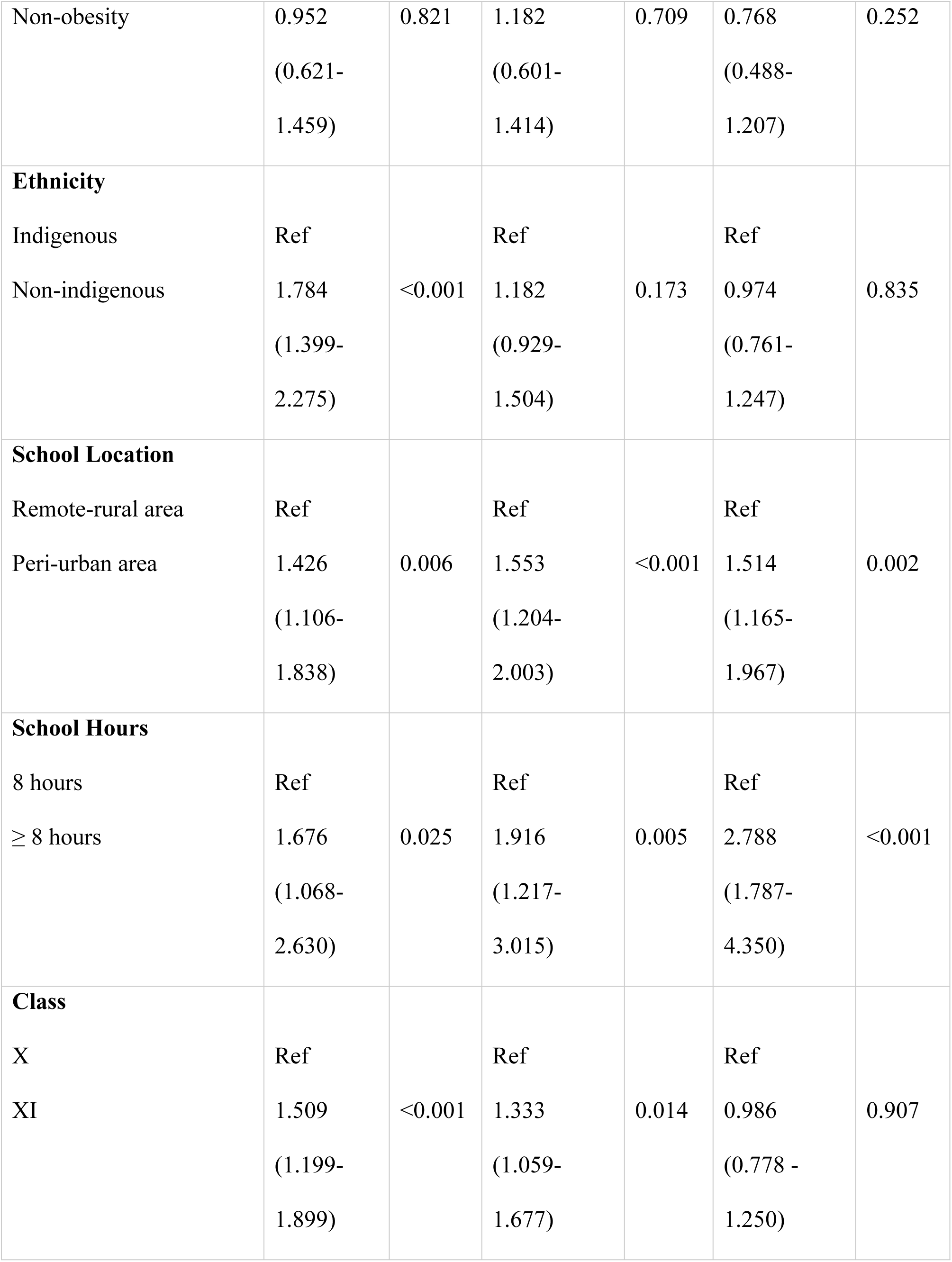
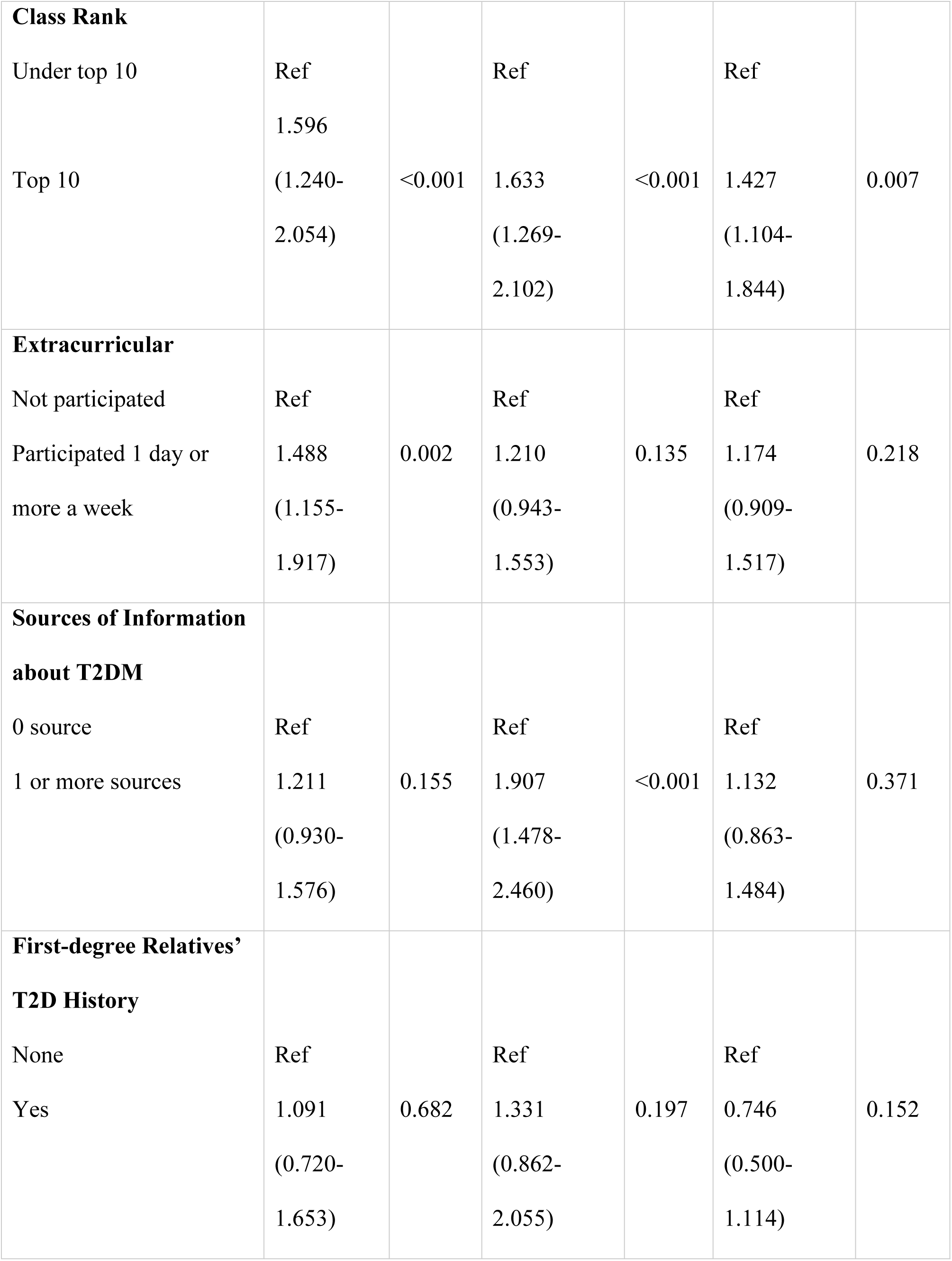
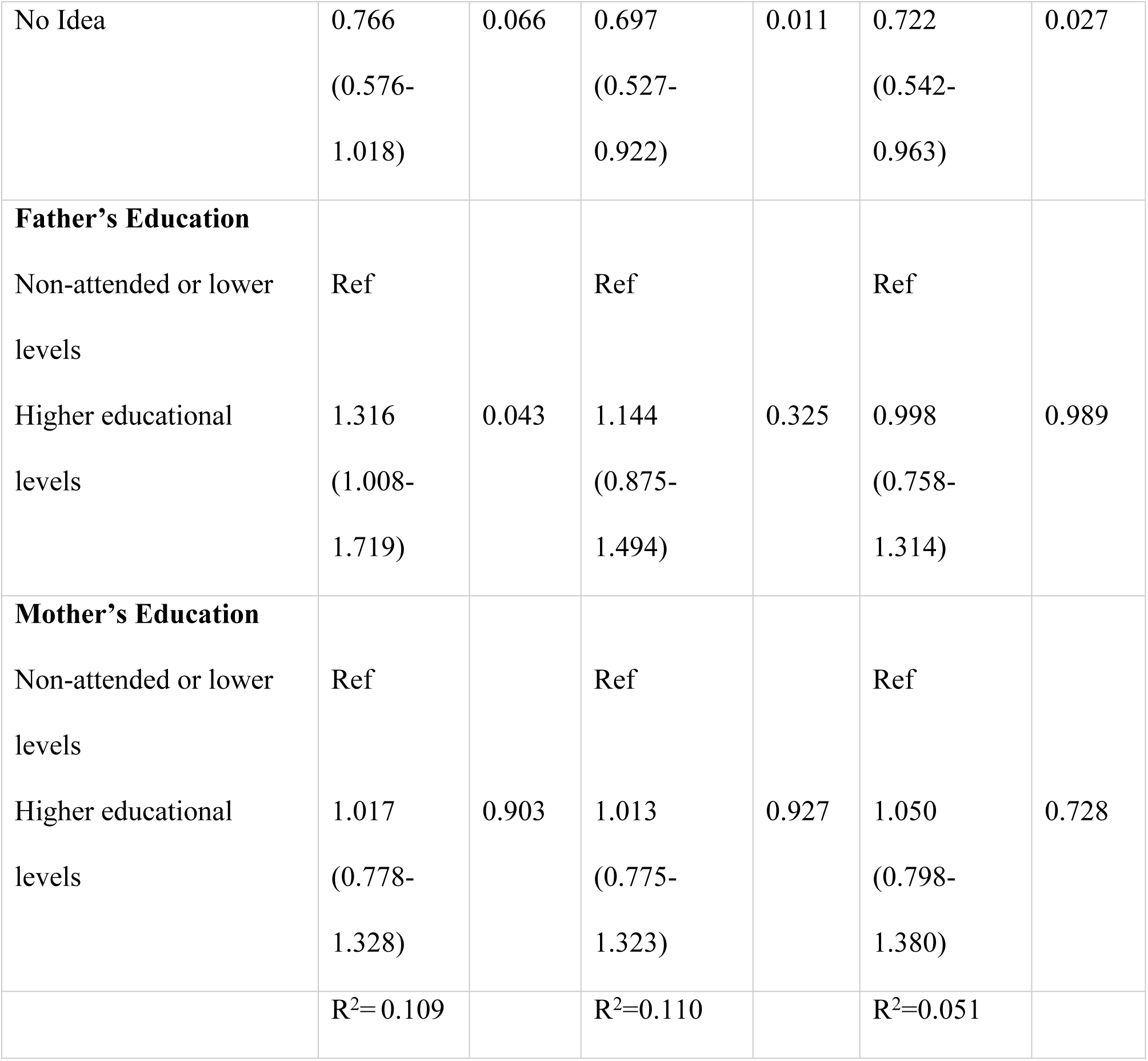
Multivariable analysis of determinants of T2DM-related KAP.

Regarding attitudes towards T2D, females (AOR = 1.542; 95% CI: 1.215–1.957), those in periurban areas (AOR = 1.553; 95% CI: 1.204–2.003), and those with longer school hours (AOR = 1.916; 95% CI: 1.217–3.015) were more likely to have positive attitudes. Students in higher grades (AOR = 1.333; 95% CI: 1.059–1.677) and those who ranked in the top ten (AOR = 1.633; 95% CI: 1.269–2.102) also demonstrated favourable attitudes. However, participants unsure of their relatives’ T2D status are less likely to have positive attitudes (AOR = 0.697; 95% CI: 0.527–0.922).

In terms of practice, adolescents in peri-urban areas and those with longer school hours demonstrated better practices, with adjusted odds ratios (AOR) of 1.514 (95% CI: 1.165–1.967) and 2.788 (95% CI: 1.787–4.350), respectively. Additionally, participants ranked in the top ten showed an AOR of 1.427 (95% CI: 1.104–1.844). Conversely, those unaware of their first-degree relatives’ T2D status were less likely to have good practices (AOR = 0.722; 95% CI: 0.542–0.963).

## Discussion

Recognizing the importance of early awareness for better identification and reduced risk, this study assessed the KAP regarding T2D among rural adolescents and identified associated sociodemographic factors. Most adolescents demonstrated a good understanding of T2D, contrasting with studies in Bangladesh, Jordan, and Nepal, where urban adolescents had better knowledge [16–17]. This discrepancy may stem from increased access to diabetes information through social media and websites, as part of national efforts to address the diabetes epidemic.

However, despite the generally encouraging level of knowledge, specific gaps were identified among students regarding T2D. While many recognized risk factors, such as family history and physical inactivity, few were aware of the risks associated with smoking and psychological stress. This pattern aligns with findings from Bangladesh, India, Kuwait, and Saudi Arabia [16, 19, 25–26]. These gaps are concerning, as they may worsen the global T2D burden. Although about a quarter mentioned receiving information from school and half from online sources, these gaps highlight the shortcomings in diabetes education, often due to a lack of integration in the curriculum, engaging teaching methods, and structured health literacy promotion [8, 16]. Additionally, in contrast to a study among adolescents in urban Jordan [18], our findings indicate that rural adolescents generally have a positive attitude toward T2D, which likely reflects their good understanding of the disease (ρ = 0.424, p < 0.001). Many believe that healthy eating and regular physical activity can help prevent T2D, consistent with findings from Bangladesh [16]. However, this positive attitude is not uniform. Many participants were uncertain or disagreed about specific risk factors, such as whether smoking increases the risk of T2D. This indicates that while adolescents generally understand T2D and healthy behaviors, they lack clarity on specific lifestyle consequences, which could undermine their preventive efforts (ρ = 0.052, p = 0.040). Such uncertainty may impede the practical application of knowledge to preventive practices [27].

Our study also found that, similar to urban adolescents in Jordan, female adolescents in rural areas had better knowledge of T2D [18]. Additionally, higher-grade-level students and top-ranking peers demonstrated greater knowledge, more positive attitudes, and better practices regarding T2DM. This finding is consistent with those from Nepal, which indicate that higher grade levels correlate with more favorable attitudes toward diabetes [17]. These characteristics suggest that these adolescents could effectively serve as peer leaders in school-based diabetes interventions. Supporting this, a study in Iran showed that peer-led approaches improved adolescents’ knowledge, dietary habits, risk awareness, disease beliefs, stress prevention, and symptom recognition [28].

Our research found that rural adolescents with more educated fathers had greater knowledge of T2D, while no correlation was observed with maternal education. This finding may reflect the patriarchal structure of the community, where fathers are seen as primary educators in health matters [29]. In contrast, a study in urban China found that adolescents with educated mothers had a better understanding of the role of free sugar in T2D [30], suggesting that maternal education influences diabetes knowledge differently across sociocultural contexts. Given that adolescents spend a significant amount of time at school [8], our findings highlight the importance of parental engagement in shaping their awareness and self-care behaviors. Notably, adolescents aware of their parents’ T2D status exhibited more positive attitudes and better self-care, underscoring the need for effective parent-child communication.

### Strengths and limitations

To our knowledge, amid the scarcity of studies on adolescents’ KAP regarding T2D, this study is the first to examine KAP levels among rural adolescents, along with associated factors, particularly in the post-pandemic context, when lifestyles, including those of adolescents in rural areas have changed. Thus, our findings provide a valuable foundation for future research on T2D prevention among youth. Additionally, we used multistage cluster sampling to represent diverse socioeconomic backgrounds across 11 of the 21 districts in Kampar Regency; however, caution is warranted regarding the generalizability of the findings. Although KAP scores were originally continuous, we categorized them using a median split to facilitate interpretation and presentation of results, particularly for public health policy makers and stakeholders. However, we acknowledge that dichotomization may lead to loss of information and reduced statistical power compared with analyses of continuous scores. participants also completed a self-administered questionnaire in a classroom setting, where peer interaction or social desirability might have shaped their responses. Finally, we did not assess adolescents’ motivation for early detection behaviors, an important indicator of good practice and a reflection of knowledge and attitudes toward the disease. This omission was partly due to the study setting, where diabetes screening was mandatory for students, thereby limiting the ability to capture voluntary health-seeking behaviors [8].

## Conclusions

This study found that adolescents in rural Indonesia generally possess good knowledge, positive attitudes, and practices regarding T2D. However, significant gaps exist in all three areas: knowledge, attitude, and practice. These gaps are particularly noticeable in adolescents’ understanding of the risk factors associated with T2D, which is essential for taking informed preventive measures. To address these gaps, structured and systematic school-based diabetes interventions should be implemented, considering significantly associated factors and actively involving parents as key partners in promoting prevention among adolescents.

## Data Availability

All relevant data are within the manuscript and its Supporting Information files

## Acknowledgements

We would like to express our sincere gratitude to Universitas Pahlawan Tuanku Tambusai, the provincial education department, and the participating schools for their support and cooperation.

## Notes

### Competing Interest Statement

The authors have declared no competing interest.

### Funding Statement

The author(s) received no specific funding for this work.

### Author Declarations

Ethical approval for the study was granted by the Institutional Review Board of Institut Kesehatan Payung Negeri (No. 277/IKES PN/KEPK/IX/2024) in Pekanbaru, Riau Province, Indonesia

